# Passive and active immunity in infants born to mothers with SARS-CoV-2 infection during pregnancy: Prospective cohort study

**DOI:** 10.1101/2021.05.01.21255871

**Authors:** Dongli Song, Mary Prahl, Stephanie L. Gaw, SudhaRani Narasimhan, Daljeet Rai, Angela Huang, Claudia Flores, Christine Y. Lin, Unurzul Jigmeddagva, Alan H.B. Wu, Lakshmi Warrier, Justine Levan, Catherine B.T. Nguyen, Perri Callaway, Lila Farrington, Gonzalo R. Acevedo, Veronica J. Gonzalez, Anna Vaaben, Phuong Nguyen, Elda Atmosfera, Constance Marleau, Christina Anderson, Sonya Misra, Monica Stemmle, Maria Cortes, Jennifer McAuley, Nicole Metz, Rupalee Patel, Matthew Nudelman, Susan Abraham, James Byrne, Priya Jegatheesan

## Abstract

**OBJECTIVE:** To investigate maternal immunoglobulins’ (IgM, IgG) response to SARS-CoV-2 infection during pregnancy and IgG transplacental transfer, to characterize neonatal antibody response to SARS-CoV-2 infection, and to longitudinally follow actively- and passively-acquired SARS-CoV-2 antibodies in infants.

**DESIGN:** A prospective observational study.

**SETTING:** A public healthcare system in Santa Clara County (CA, USA).

**PARTICIPANTS:** Women with SARS-CoV-2 infection during pregnancy and their infants were enrolled between April 15, 2020 and March 31, 2021.

**OUTCOMES:** SARS-CoV-2 serology analyses in the cord and maternal blood at delivery and longitudinally in infant blood between birth and 28 weeks of life.

**RESULTS:** Of 145 mothers who tested positive for SARS-CoV-2 during pregnancy, 86 had symptomatic infections: 78 with mild-moderate symptoms, and eight with severe-critical symptoms. Of the 147 newborns, two infants showed seroconversion at two weeks of age with high levels of IgM and IgG, including one premature infant with confirmed intrapartum infection. The seropositivity rates of the mothers at delivery was 65% (95% CI 0.56-0.73) and the cord blood was 58% (95% CI 0.49-0.66). IgG levels significantly correlated between the maternal and cord blood (Rs= 0.93, p< 0.0001). IgG transplacental transfer ratio was significantly higher when the first maternal positive PCR was 60-180 days before delivery compared to <60 days (1.2 vs. 0.6, p=<0.0001). Infant IgG negative conversion rate over follow-up periods of 1-4, 5-12, and 13-28 weeks were 8% (4/48), 12% (3/25), and 38% (5/13), respectively. The IgG seropositivity in the infants was positively related to IgG levels in the cord blood and persisted up to six months of age.

**CONCLUSIONS:** Maternal SARS-CoV-2 IgG is efficiently transferred across the placenta when infections occur more than two months before delivery. Maternally-derived passive immunity may protect infants up to six months of life. Neonates mount a strong antibody response to perinatal SARS-CoV-2 infection.

## Introduction

An important aspect of immunity against infectious pathogens in young infants relies on effective maternal antibody production, transfer of maternal antibodies across the placenta to the fetus, and persistence of passive immunity in the infant. Our understanding of the immune response to severe acute respiratory syndrome coronavirus-2 (SARS-CoV-2) is expanding rapidly through extensive basic and clinical studies.^1-4^ However, the literature on SARS-CoV-2 immunity in pregnant mothers and infants remains limited.^5-9^ Global efforts are focused on controlling the COVID-19 pandemic through public health prevention measures and universal vaccination. Knowledge of neonatal immune response to SARS-CoV-2 and maternally-derived passive immunity in young infants is urgently needed to guide ongoing COVID-19 infection prevention and vaccination strategies in pregnant mothers and infants.

Recent publications have shown evidence of maternal SARS-CoV-2 antibody transplacental transfer.^6,7,9^ However, the majority of maternal SARS-CoV-2 infections in these reports occurred late in pregnancy, as these studies were conducted during the first few months of the COVID-19 pandemic. Therefore, the timing and efficiency of maternal antibody production and transplacental transfer throughout gestation remain to be fully understood, which has important implications for the timing of maternal immunization to benefit both pregnant mothers and their young infants. Furthermore, the important question as to the persistence of maternally-derived passive immunity in infants needs to be investigated. While SARS-CoV-2 infection has been described in newborns,^10,11^ little is known about infant immune response to perinatal infection. The aims of this study were to investigate SARS-CoV-2 antibody transplacental transfer with respect to the timing of maternal infection during gestation, antibody response to SARS-CoV-2 infection in the newborns, and persistence of passively- and actively-acquired SARS-CoV-2 antibodies in infants.

## Methods

### Study design, participants, and procedures

This is a prospective observational study of pregnant mothers with SARS-CoV-2 infection during pregnancy and their infants. The study was conducted from April 15, 2020 to March 31, 2021, in a public healthcare system, including one regional medical center and two community hospitals. The healthcare system primarily serves the medically indigent population of Santa Clara County California (USA). The study was approved by the institutional review boards of Santa Clara Valley Medical Center. Patients or the public were not involved in the design, or conduct, or reporting, or dissemination plans of our research.

In April 2020, our health system implemented a universal screening protocol for SARS-CoV-2 infection in women presenting in labor or within three days prior to admission for elective deliveries.^12^ SARS-CoV-2 infection was diagnosed based on a positive SARS-CoV-2 reverse transcriptase polymerase chain reaction (RT-PCR) test using nasopharyngeal specimen performed either before delivery or through universal screening at delivery. The timing of maternal SARS-CoV-2 infection was based on the first positive SARS-CoV-2 PCR test. The severity of SARS-CoV-2 symptoms (mild, moderate, severe, or critical) was assessed according to the Society for Maternal-Fetal Medicine guidelines.^13^ If the maternal infection was within 10-14 days of delivery, the mother and infant roomed in together with airborne isolation precautions and the mother wore a surgical mask when holding and breastfeeding the baby during the isolation period. The nasopharyngeal SARS-CoV-2 PCR was performed in the newborns at 24 hours of life. The infants were retested between 48-72 hours of life if they were in the Neonatal Intensive Care Unit (NICU).

Maternal and cord blood were collected at the time of delivery. Serial infant blood samples were collected between the ages of 1-28 weeks, coordinated with their clinic visits. Levels of SARS-CoV-2 immunoglobulin M (IgM) and immunoglobulin G (IgG) to the spike protein receptor binding domain (RBD) and nucleocapsid protein (NP) of SARS-CoV-2 were measured using the Pylon 3D automated immunoassay system (ET Healthcare, Palo Alto, CA) as previously described.^14^ The background corrected signal was reported as relative fluorescent units (RFU), which is proportional to the amount of specific antibodies in the sample allowing for quantification. The positive cutoffs for IgM and IgG were set to >50 RFU to achieve 100% specificity and a high level of sensitivity.^14^ Quantitative reverse transcriptase PCR (qRT-PCR) was performed on maternal blood, cord blood, placenta, and meconium in a subset of infants. Primer sequences targeted the N and Orf1b SARS-CoV-2 genes (supplemental Methods, supplemental Tables 1 and 2).

**Table 1.** Maternal and neonatal demographics and outcomes.

|  | **Maternal and infant serology cohort** |
| --- | --- |
| Mothers, n | 145 |
| Newborns, n | 147 |
| **Maternal demographics and outcomes** |  |
| Maternal age, years, median (range) | 27 (16, 42) |
| Gravida, median (range) | 3 (1, 12) |
| Para, median (range) | 1 (0, 9) |
| Hispanic, n (%) | 126 (87) |
| Race, n (%) |  |
| White, n (%) | 130 (90) |
| Black, n (%) | 6 (4) |
| Asian, n (%) | 9 (6) |
| Asymptomatic, n (%) | 59 (41) |
| Mild to moderately symptomatic, n (%) | 78 (54) |
| Severe to critically symptomatic, n (%) | 8 (6) |
| Symptomatic at the time of diagnosis, n (%) | 86 (59) |
| Symptomatic at the time of delivery, n (%) | 22 (15) |
| Cesarean section, n (%) | 46 (32) |
| Multiple pregnancies, n (%) | 3 (2) |
| Maternal diabetes, n (%) | 29 (20) |
| Maternal hypertension, n (%) | 30 (21) |
| Maternal obesity, n (%) | 33 (23) |
| Preterm delivery, n (%) | 15 (10) |
| Intrauterine fetal demise, n (%) | 1 (1) |
| **Neonatal demographics and outcomes** |  |
| Gestational age, weeks, median (range) | 39.1 (27.4, 41.6) |
| Birth weight, grams, median (range) | 3285 (990, 4670) |
| Breastfeeding in the hospital, n (%) | 143 (97) |
| Exclusive breastfeeding in the hospital, n (%) | 85 (58) |
| Rooming in with mother, n (%) | 132 (90) |
| NICU admission, n (%) <sup>a</sup> | 23 (16) |
| Length of stay during birth hospital, days, median (range) | 2 (1, 81) |
| SARS-CoV-2 nasopharyngeal swab positive, n (%) <sup>b</sup> | 1 (1) |

<sup>a</sup>Reasons for NICU admissions: 7 for prematurity, 1 for a congenital anomaly, 1 for dehydration and 14 for respiratory distress, metabolic acidosis and or evaluation for infection.
<sup>b</sup>SARS-COV-2 PCR using nasopharyngeal specimens was performed in 70 (99%) of the newborns born to mothers who were first PCR positive within two weeks of delivery.

**Table 2.** Maternal and cord blood serology and timing of maternal first positive PCR.

|  | **Total** | **2w**<br>**0-13d** | **2w-2m**<br>**14-59d** | **2m-6m**<br>**60-179d** | **>6m**<br>**≥180d** |
| --- | --- | --- | --- | --- | --- |
| **Maternal serology, n** | **129** | **56** | **28** | **36** | **9** |
| IgM- and IgG-, n | 45 | 35 | 4 | 5 | 1 |
| IgM+ and IgG-, n | 4 | 4 | 0 | 0 | 0 |
| IgM+ and IgG+, n | 29 | 6 | 13 | 9<sup>a</sup> | 1<sup>b</sup> |
| IgM- and IgG+, n | 51 | 11 | 11 | 22 | 7 |
| IgM+ and/or IgG+, n | 84 | 21 | 24 | 31 | 8 |
| IgM, RFU, median (range) | 27 (7, 1388) | 25.5 (2, 315) | 34.5 (7, 1388) | 26.5 (11, 263) | 25 (7, 59) |
| IgG, RFU, median (range) | 84 (1, 3582) | 22.5 (1, 401) | 178 (1, 1123) | 194.5 (22, 2311) | 199 (41, 3582) |
| **Cord blood serology, n** | **144** | **70** | **27** | **38** | **9** |
| IgG-, n | 61 | 48 | 8 | 4 | 1 |
| IgG+, n | 83 | 22 | 18 | 32 | 8 |
| IgG, RFU, median (range) | 66.5 (0, 2916) | 14 (0, 1820) | 77 (2, 1164) | 232 (22, 2916) | 209 (45, 1173) |
| **Paired cord and maternal blood serology, n** | **125** | **54** | **26** | **36** | **9** |
| Maternal IgG + and Cord blood IgG +, n | 69 | 12 | 19 | 31 | 7 |
| Maternal IgG + and cord blood IgG -, n | 8 | 4 | 3 | 0 | 1 |
| Maternal IgG – and cord blood IgG -, n | 45 | 37 | 4 | 4 | 0 |
| Maternal IgG – and cord blood IgG, +, n | 3 | 1 | 0 | 1 | 1 |

RFU=Relative fluorescent unit.
<sup>a</sup> All 9 mothers' first positive SARS-VoC-2 PCR were between 63 and 103 days before delivery.
<sup>b</sup> This mother's SARS-VoC-2 PCR was positive at 10 weeks gestation and was positive again at the time of delivery at 39 weeks gestation.

### Data collection and analysis

Clinical data included maternal and neonatal demographics, the severity of maternal symptoms of SARS-CoV-2 infection, days between maternal first positive SARS-CoV-2 PCR test and delivery, and neonatal outcomes. Demographics, clinical outcomes, and serum IgM and IgG levels were summarized using descriptive analyses. Transplacental IgG transfer ratios were calculated by dividing cord blood IgG levels by maternal blood IgG levels. Correlation between maternal and cord blood IgG levels and correlation between placental transfer ratio and gestational age (GA) at birth were analyzed using Spearman’s rank-order correlation. The transfer ratios were compared between maternal groups based on infection severity and time between first maternal positive PCR and delivery using the Kruskal-Wallis test, followed by Dunn’s test for pairwise multiple comparisons with the Holm-Sidák stepwise adjustment.

## Results

During the study period, 3936 mothers delivered in the health system with 3956 live births, and 254 (6.5%) of the mothers had at least one positive SARS-CoV-2 PCR test during the pregnancy. The study enrolled 145 mothers with SARS-CoV-2 infection and 147 of their infants (Figure 1). Of 145 enrolled mothers, 86 (59%) had symptomatic infection, including 78 with mild-moderate symptoms and eight with severe-critical symptoms (Table 1). The distribution of the severity of the maternal infection is shown in supplemental Table 3. Of 147 newborns, 23 (16%) were admitted to the NICU. SARS-CoV-2 PCR was performed on nasopharyngeal specimens of 89 newborns at 24 hours of life, and only one 31-week preterm infant tested positive.

**Figure 1:**
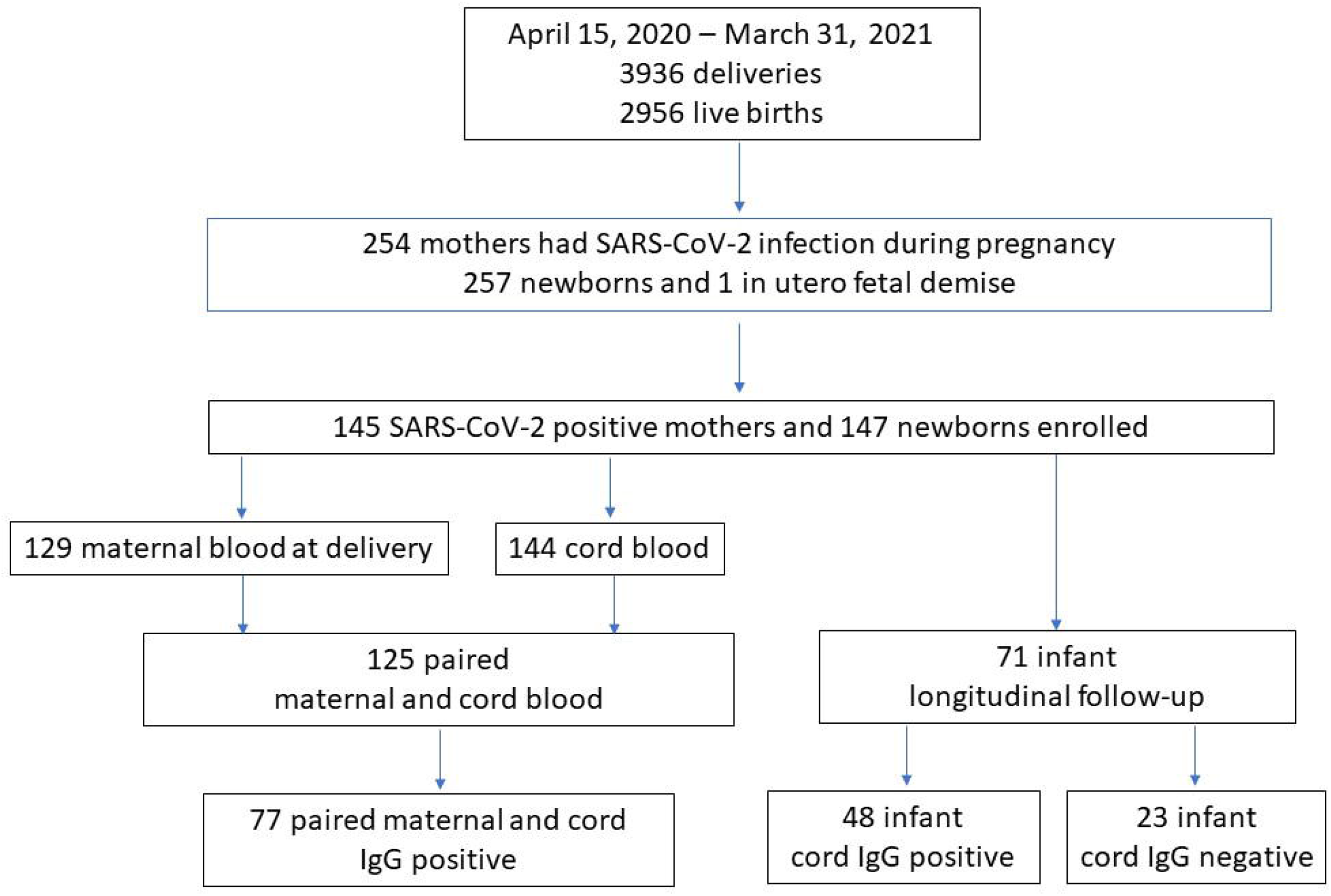
Study participants enrollment.

### Maternal and cord blood serology

Serum serology was performed on 129 mothers at delivery and 144 cord blood samples. The temporal profiles of maternal and cord blood IgM and IgG with respect to the timing of first maternal PCR positivity are shown in Figure 2. Antibody status and levels in maternal and cord blood were evaluated in four groups based on the days between maternal first positive SARS-CoV-2 PCR and delivery (<14 days, 14 to 59 days, 60 to 180 days, and >180 days) (Table 2). The maternal seropositivity rate at delivery was 65% (84/129, 95% CI 0.56-0.73), and the cord blood IgG positivity rate was 58% (83/144, 95% CI 0.49-0.66).

**Figure 2:**
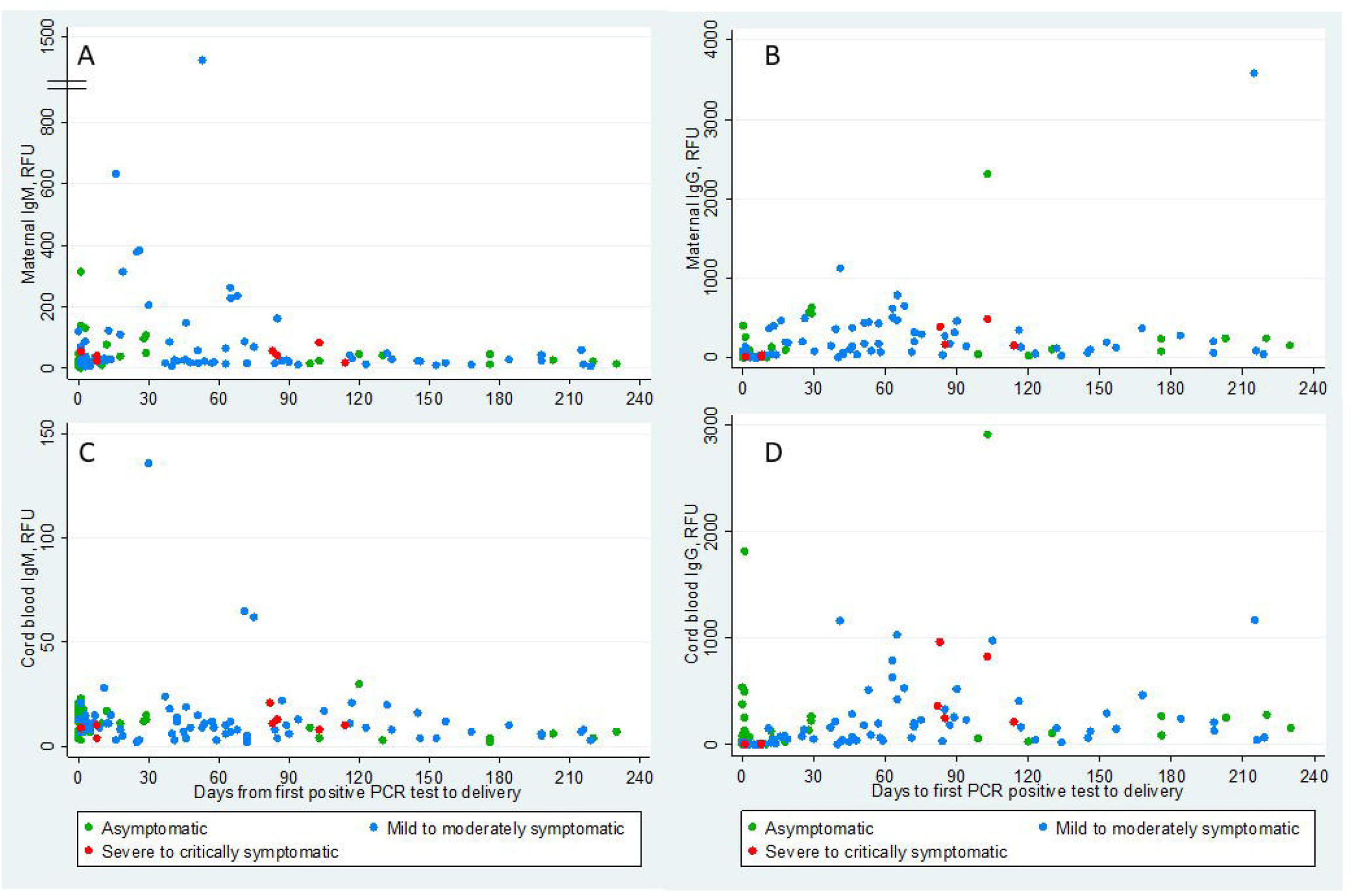
Temporal distribution of maternal and cord blood IgM and IgG. Panel A, B. scatterplots show the distribution of maternal blood SARS-CoV-2 IgM and IgG levels in relative fluorescent unit (RFU) at the time of delivery in Y-axis and days from maternal first positive SARS-CoV-2 PCR test to delivery in X-axis. Panel C, D scatterplots show the distribution of cord blood SARS-CoV-2 IgM and IgG levels in RFU at the time of delivery in Y-axis and days from maternal first positive SARS-CoV-2 PCR test to delivery in X-axis. The different colors represent the severity of the maternal symptoms at the time of diagnosis.

Paired serology analysis was performed in 125 maternal-cord blood samples (Table 2). Of the 77 IgG positive mothers, 69 (90%) of their newborns’ cord sera were positive for IgG. Of the eight IgG negative infants, seven were born to mothers with infection within 45 days of delivery, and one was born to a mother who had a positive PCR at 254 days before delivery. Of the 48 IgG negative mothers, 45 (94%) of their newborns’ cord sera were negative for IgG. Notably, three infants whose cord blood was positive for IgM (65, 136, and 62 RFU) were born to mothers whose blood was also positive for IgM at the time of delivery. The follow-up serology tests for two of the infants at two and three weeks of age were negative for IgM and IgG. No follow-up serology was available for the third infant. Available delivery specimens (maternal and cord blood, placenta, and meconium) were evaluated by SARS-CoV-2 PCR and found to be negative for all three infants (supplemental Table 4).

There was a significant positive correlation between IgG levels in the 125 paired maternal and cord blood samples (Rs=0.93, p<0·0001, Figure 3A). Transplacental IgG transfer ratios were calculated in 77 IgG positive mother-infant dyads, and the median transfer ratio was 1.0 (95% CI 0.86-1.09). The transfer ratio was significantly higher in the mothers who were severe-critically symptomatic (n=4) compared to mothers who were asymptomatic (n=23) (1.6 vs. 1.0, p=0.003) or mild-moderately symptomatic (n=50) (1.6 vs. 0.9, p=0.002). To illustrate the temporal effect of maternal infection on transfer efficiency, we analyzed transfer ratios of 54 symptomatic mother-infant dyads. Asymptomatic mothers were excluded from this analysis as their timing of infections cannot be concluded definitively from the timing of PCR positivity (Figure 3B). The transfer ratios based on time elapsed from the first maternal positive PCR to delivery were 0.6 (<60 days, n=22), 1.2 (60-180 days, n=27), and 0.9 (>180 days, n=5). The ratio was significantly higher in the 60-180 days group compared to the <60 days group (1.2 vs. 0.6, p=<0.0001). Transfer ratios based on the trimester of maternal infection were 0.9 (1st trimester, n=7), 1.2 (2nd trimester, n=9), and 0.9 (3rd trimester, n=38) (Figure 3D). The ratio was significantly higher in second trimester infections than third trimester infections (1.2 vs. 0.9, p=0.02). There was no significant correlation between the transfer ratio and GA at birth (Rs=0.18, p=0.1, Figure 3C); however, 95% of the infants in our cohort were born at greater than 34 weeks gestation.

**Figure 3:**
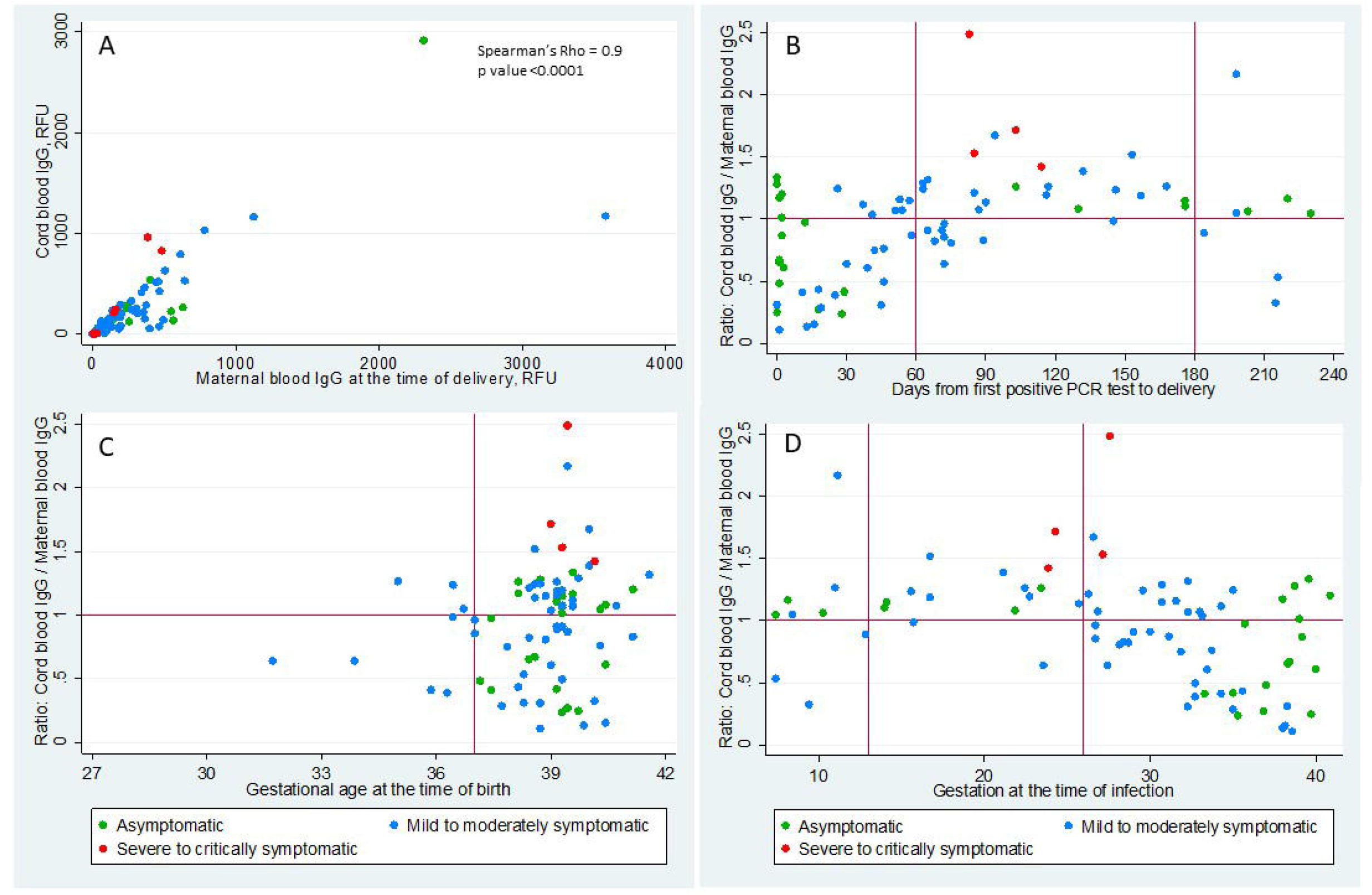
Correlation of cord blood and maternal IgG and distribution of IgG transplacental transfer ratio. Panel A. Scatterplot shows the correlation of cord blood SARS-CoV-2 IgG levels in Y-axis and maternal blood SARS-CoV-2 IgG levels in X-axis in relative fluorescent unit (RFU). Panel B. Scatterplot shows the distribution of IgG transplacental ratio (cord blood/maternal blood SARS-CoV-2 IgG levels) in the Y-axis and days from maternal first positive SARS-CoV-2 PCR test to delivery in X-axis. Panel C. Scatterplot shows the distribution of IgG transplacental ratio in the Y-axis and gestational age at the time of delivery in X-axis. Panel D. Scatterplot shows the distribution of IgG transplacental ratio in the Y-axis and gestational age at the time of maternal first positive SARS-CoV-2 PCR test in X-axis. The different colors represent the severity of the maternal symptoms at the time of diagnosis.

### Maternally-derived IgG longitudinal follow-up in infants

To evaluate maternally-derived IgG persistence postnatally, we followed serology in 48 infants with positive cord IgG. All infants showed a steady decrease in IgG levels over time (Figure 4A). The IgG seroconversion rate was calculated for those infants who had at least one serology test during the follow-up age periods of 1-4 weeks, 5-12 weeks, and 13-28 weeks. The negative IgG conversion rates for the three follow-up periods were 8% (4/48), 12% (3/25), and 38% (5/13), respectively. The infants who had lower levels of IgG in the cord blood became IgG negative earlier; the cord IgG levels of those infants who seroconverted during the three follow-up periods ranged between 52-66 RFU, 68-150 RFU, and 123-251 RFU, respectively. Two infants who had cord IgG levels greater than 500 RFU remained seropositive at 27 weeks of age.

**Figure 4:**
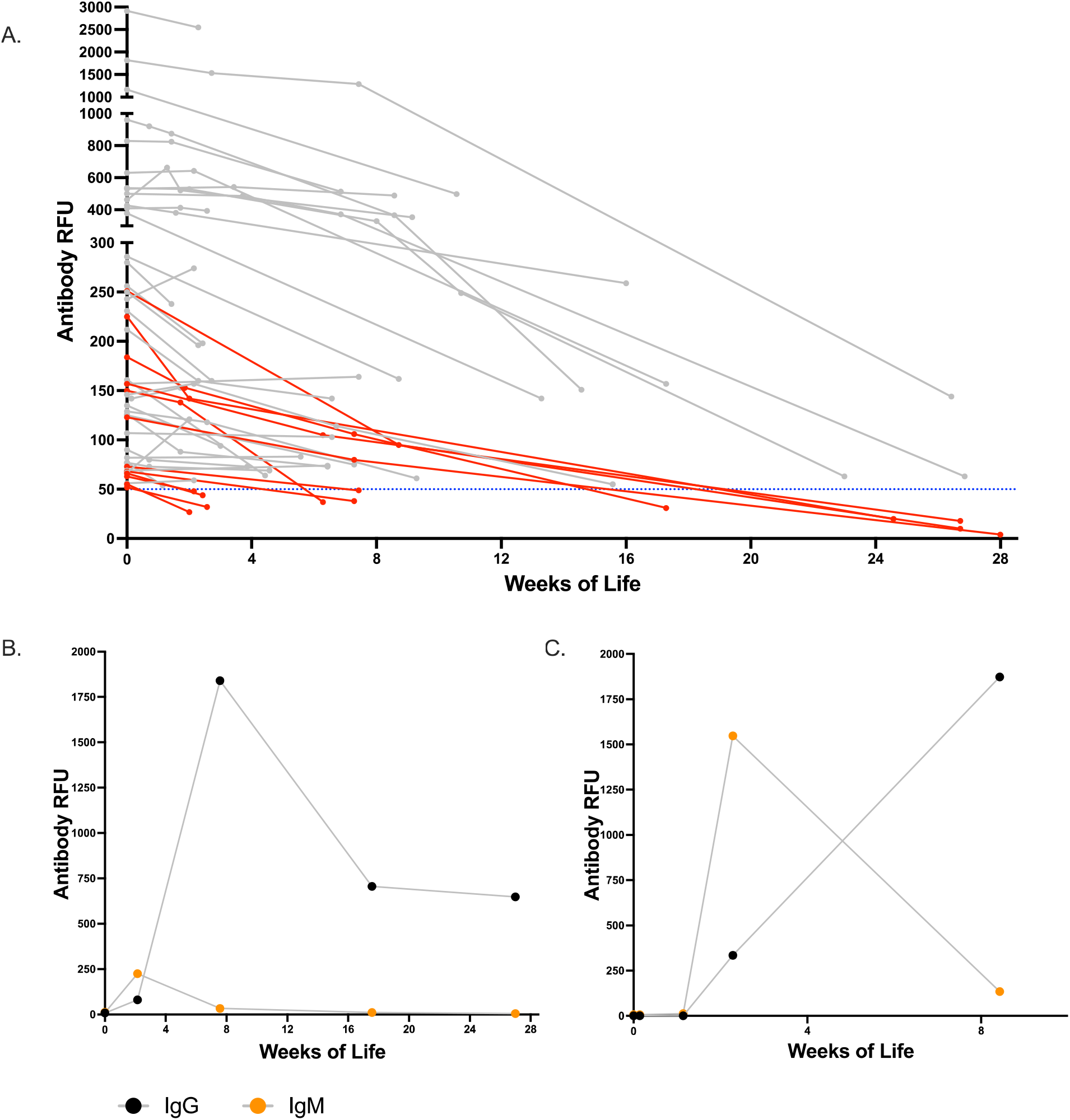
Longitudinal follow-up of SARS-CoV-2 antibody levels in infants. Panel A shows the longitudinal IgG levels of the infants who had cord blood IgG level >50 relative fluorescent unit (RFU). The infants’ IgG levels in RFU is shown in Y-axis, and the age of the infant in weeks at the time of follow-up is shown in X-axis. The infants whose IgG became negative, <50RFU, during the longitudinal follow up are shown in red color. Panel B shows the IgG and IgM levels of the term infant whose cord antibody was negative and seroconverted at 2 weeks of life. Panel C shows the IgG and IgM levels of the 31 weeks preterm infant with confirmed intrapartum SARS-CoV-2 infection whose cord antibody was negative and seroconverted at 2 weeks of life.

### Infant antibody response to perinatal SARS-CoV-2 infection

We performed surveillance serology tests at 2-4 weeks of age in 23 of 41 (56%) infants who had negative serology in the cord blood and were born to mothers with first positive PCR <14 days before delivery.

Two infants showed seroconversion, including the 31-week preterm infant who tested positive for SARS-CoV-2 and a term infant. Interestingly, both infants were born to asymptomatic mothers who tested positive for SARS-CoV-2 PCR for the first time at delivery and negative for SARS-CoV-2 antibodies, indicating a new onset of infection. Both infants were asymptomatic for SARS-CoV-2 infection. The preterm infant, born to a mother with spontaneous preterm labor, was admitted to the NICU immediately after birth, isolated from the mother for 14 days, and discharged home at 17 days of life after an unremarkable NICU course. The infant’s cord blood SARS-VoC-2 PCR was negative, but nasopharyngeal PCR was positive at 24 hours of life and remained positive at discharge. Additionally, the infant’s meconium and maternal blood at the time of delivery were PCR positive. The term infant roomed in with the mother in the postpartum unit and was discharged home at two days of life. This infants’ cord blood and nasopharyngeal SARS-CoV-2 PCR were negative at 24 hours of life, and nasopharyngeal PCR was not repeated.

The preterm infant showed serial negative serology tests after birth on days two, four, and eight, then seroconverted on day 16 (IgM 1548 RFU, IgG 335 RFU) (Figure 4B). The infant’s IgM decreased to 134 RFU, and IgG increased to 1873 RFU at eight weeks. The term infant had the first follow-up test at two weeks and was found positive for IgM (225 RFU) and IgG (80RFU) (Figure 4C). The infant’s IgM became negative, and IgG peaked at 1841 RFU at eight weeks; the IgG subsequently decreased to 648 RFU at 24 weeks.

## Discussion

We conducted a prospective observational study in 145 pregnant mothers with SARS-CoV-2 infections during pregnancy and 147 of their infants. The majority of infected mothers seroconverted before delivery. The IgG levels in maternal blood at delivery and cord blood were highly correlated. High transplacental IgG transfer ratios were observed when infection onset was greater than 60 days prior to delivery or in the second trimester. The persistence of maternal-derived IgG in infants was positively correlated to the initial cord blood level. Additionally, we showed strong antibody responses to perinatal SARS-CoV-2 infection in two asymptomatic neonates.

In our study, 6.5% of mothers presenting for delivery had at least one positive SARS-CoV-2 PCR during their current pregnancy. The majority of mothers had asymptomatic or mild-moderate infections, consistent with previous cohort studies.^15,16^ The maternal IgG levels at delivery were relatively low, comparable to levels in non-ICU patients.^14^ Importantly, the temporal profiles of maternal and cord blood IgG levels were in parallel, peaking around 60-90 days post maternal infection. The timing and efficiency of maternal IgG transfer have important implications for developing maternal immunization strategies to protect infants.^17-19^ For example, in maternal pertussis immunization, infant seropositivity rate and cord blood IgG levels to pertussis toxin were higher following Tdap immunization during the second trimester than during the third trimester. We studied pregnant mothers who had SARS-CoV-2 infections in all three trimesters and provide a comprehensive profile of transplacental IgG transfer with respect to the timing of infections throughout gestation. We observed that transfer ratio was 0.6 when infection onset was less than 60 days before delivery; plateaus at 1.2 and 0.9 when infections occurred 60-180 days and greater than 180 days before delivery. Prior studies of pregnancy related infection in the last 70 days of gestation found impaired SARS-CoV-2 IgG transplacental transfer (ratio 0.7).^7,8^ Another study characterized a cohort of pregnant mothers who had infections during the last 120 days of gestation and showed that transfer ratios increased with length of time from infection to delivery, with transfer ratios reached above 1.0 in the majority of mothers.^6^ Taken together, these studies demonstrate that cross-placental SARS-CoV-2 IgG transfer occurs throughout gestation, and a higher transfer efficiency is achieved when infection onset is more than two months prior to delivery. Matching the peak IgG transplacental transfer and the peak immune response after maternal infection may result in high cord IgG. Information from these maternal and cord serology studies is important for guiding the timing of maternal vaccination in pregnancy to optimize neonatal immunity in concert.

In our study, the persistence of maternal-derived IgG in infants showed a wide range, from two weeks to more than 26 weeks of age. An important observation is that IgG positivity in infants is positively associated with the initial cord IgG levels that are determined by maternal IgG levels and transplacental transfer efficiency. As more pregnant mothers are vaccinated for SARS-CoV-2, knowledge of passive immunity in infants may inform mother-infant care and SARS-CoV-2 vaccination strategy in infants.

Consistent with prior literature showing rare vertical maternal-fetal transmission,^20-23^ we found only one infant with confirmed intrapartum acquired neonatal infection. (21) This infant was seronegative in cord blood and during the first week of life but seroconverted at two weeks of life, providing insight into the timing of infant seroconversion in the setting of intrapartum infection. We identified another infant who seroconverted at two weeks follow-up test; however, available virology and serology data is not sufficient to determine the timing and mode of this perinatal infection. Clinical presentations of perinatal SARS-CoV-2 infection have been described previously;^10,11 24^ however, little is known about neonatal serology response and long-term clinical outcomes. Interestingly, both infants in our study had asymptomatic infection but mounted strong antibody responses; the timing of seroconversion and levels of IgM and IgG are comparable to that observed in adult patients with severe disease.^14^ Both infants remained asymptomatic in the first months of life. Their long-term clinical outcomes, along with immune status, will be followed. Additionally, these two cases highlight the increased risk for perinatal SARS-CoV-2 infection in infants born to mothers who have new-onset infections around the time of delivery,^10^ with implications for developing targeted protection measures and postnatal antibody screening for high-risk newborns. In our study, three infants were positive for IgM in cord blood but negative for SARS-CoV-2 virologically in birth specimens and negative for IgM and IgG at two and three weeks of age, suggesting these transient IgM levels may be false positives or maternal blood contamination. There were two prior case reports describing similar transient positive IgM levels in the cord blood without virological evidence of infection.^25,26^ Thus, diagnosis of congenital SARS-CoV-2 infection cannot be made based solely on the presence of IgM in the cord blood.^27-30^

This maternal-infant serology study, one of the largest cohorts to date, included pregnant mothers with SARS-CoV-2 infection in all three trimesters of pregnancy and provided a more comprehensive understanding of maternal SARS-CoV-2 IgG transplacental transfer. This is the first longitudinal study that has followed the level of maternally-derived SARS-CoV-2 IgG in infants up to 28 weeks and neonatal serology response after perinatal SARS-CoV-2 infection up to 24 weeks. Another strength of the study is that the cohort is representative of COVID-19 in the community. Over 90% of the mothers in this cohort are Hispanic, a population highly impacted by the COVID-19 pandemic. Our study has several limitations. It was conducted in a single healthcare system. The timing of infection was based on the first positive PCR, which might not represent the precise timing of infection in asymptomatic mothers. Our cohort had few severe cases and premature births before 35 weeks of gestation. Universal screening at the time of admission also introduces a bias in the identification of asymptomatic SARS-CoV-2 cases at or near-term gestation, as the universal screening was not implemented in our prenatal care visits and asymptomatic screening was not readily available in our general community during the study period.

## Conclusion

Our study provides insights into the intricate connections between the timing of maternal SARS-CoV-2 infection, dynamics of maternal antibody production, and transplacental immunity transfer. These processes determine the level of maternally-derived IgG in infants at birth, which in turn affects persistence of passive immunity in infants. Neonates are capable of mounting strong serology responses to perinatal SARS-CoV-2 infection. These findings have important implications in determining optimal timing of vaccination in pregnant mothers and infants. Future investigations are needed to address the durability and protection of passively and actively acquired antibodies in the infant.

## Data Availability

De-identified data is published in Mendeley data sharing site and available in the following doi:10.17632/6scfwt55fd.2.

http://dx.doi.org/10.17632/6scfwt55fd.2

## Acknowledgements

Thank you to Dr. Margaret Feeney for support of these experiments and Robin D. Wu who reviewed the manuscript and provided helpful comments and suggestions. We thank the mothers, newborns and their families who participated in the study; staff and providers in Labor and Delivery, Post-partum unit, NICU, pathology and laboratory services, at Santa Clara Valley Medical Center and O’Connor Hospital; outpatient pediatrics clinic, and BRIDGE home follow up program. We thank First 5 of Santa Clara County for their support.

## Strengths and limitations of this study

- This study included pregnant mothers with SARS-CoV-2 infection in all three trimesters of pregnancy and provided a comprehensive understanding of maternal SARS-CoV-2 IgG transplacental transfer throughout pregnancy.
- This is the first longitudinal study that has followed maternally-derived SARS-CoV-2 IgG in infants up to 28 weeks.
- This is the first study, to our knowledge, that characterized neonatal serology response to perinatal SARS-CoV-2.
- In asymptomatic mothers the first positive PCR might not represent the precise timing of infection.
- The cohort had few severe cases of maternal infection and premature births before 35 weeks of gestation.

## Author contributions

D.S. conceptualized and designed the project, participated in patient enrollment, sample collection, data visualization and interpretation, and wrote the manuscript draft.

P.J. conceptualized and designed the project, participated in patient enrollment, sample collection, data collection, analysis, interpretation, and edited manuscript.

M.P., S.L.G designed collection and processing protocols, performed sample processing, oversaw experiments and data analysis, provided funding, edited the manuscript.

S.R.N. conceptualized and designed the project, participated in patient enrollment, sample collection, data collection and edited manuscript.

D.R. participated in study design, patient enrollment, and sample collection.

A.H. coordinated data collection and management, participated in sample collection, and oversaw the implementation of the project.

C.V.F. participated in patient enrollment, coordinated sample collections, processing, and data management

L.W. designed collection and processing protocols, performed sample processing and data collection.

J.L. and C.B.T.N., CYL, UJ, VJG designed and performed experiments, performed sample processing and data collection.

P.C., L.F, G.R.A., A.V. optimized collection and processing protocols, performed sample processing and data collection

A.H.B.W. designed and oversaw serology assays.

E.A., P.N., C.M. oversaw sample collections and processing.

C.A., S.M., M.S., M.C., J.M., S.A., N.M. participated in patient enrollment and sample collection

M.N. participated in data analysis and preparing the figures for the manuscript.

R.P. participated in the study implementation, and sample collection.

J.B. designed, and oversaw the patient recruitment and implementation of the project All authors reviewed and approved the manuscript.

## Supplemental Materials

### PCR Methods

#### RNA Extraction

Maternal blood, cord blood, placental tissue, and infant meconium RNA was extracted using the QIAmp Viral RNA Mini Kit following the manufacturer’s instructions with some adjustments. 300uL of maternal and cord blood in RNAlater (1:1.3 ratio) were used for each extraction. 15-25 mg of placenta and 300µg of meconium in viral transport media was used for extraction. The kit protocol was followed with buffer amounts scaled up proportionally for the starting amount. RNA was eluted in a 40uL elution buffer for blood and 20uL elution buffer for placenta and meconium. RNA quantity was measured using the Qubit RNA High Sensitivity Assay Kit.

#### Quantitative real-time PCR

Quantitative polymerase chain reaction was performed using the ABI StepOne Plus system. Primer sequences targeted the N (nucleotide) and Orf1b (ORF1b-nsp14) gene. Primer sequences are as follows: forward primer targeting N gene \ (HKU-NF): 5’-TAATCAGACAAGGAACTGATTA-3’;Reverse primer (HKU-NR): 5’-CGAAGGTGTGACTTCCATG-3’; and Probe (HKU-NP): 5’-FAM-GCAAATTGTGCAATTTGCGG-TAMRA-3’. Forward primer targeting Orf1b-nsp14 gene (HKU-ORF1b-nsp14F): 5’-TGGGGYTTTACRGGTAACCT-3’; Reverse primer (HKU-ORF1b-nsp14R): 5’-AACRCGCTTAACAAAGCACTC-3’; and Probe (HKU-ORF1b-nsp141P): 5’-FAM-TAGTTGTGATGCWATCATGACTAG-TAMRA-3. RT-qPCR reactions were performed using the TaqMan Fast Virus 1-step Master Mix according to the manufacturer’s instructions.

**Supplemental Table 1:** PCR Reagents.

| Reagent | Volume per rxn (µL) |
| --- | --- |
| Water (RNase free) | 7.5 |
| TaqMan Fast Virus 1-step (4X) | 5 |
| Forward Primer (10 µM) | 1 |
| Reverse Primer (10 µM) | 1 |
| Probe (10 µM) | 0.5 |
| RNA Sample | 5 |

**Supplemental Table 2:** PCR Cycle.

| Steps | Temperature (C) | Time (mm:ss) # cycles |
| --- | --- | --- |
| Reverse Transcription | 50 | 05:00 |
| RT Inactivation/denaturation | 96 | 00:20 |
| Amplification | 95 | 00:05:40 |
| Amplification | 60 | 00:30 |

**Supplemental Table 3:** Distribution of severity of maternal symptoms at the time of diagnosis.

|  | **Asymptomatic**<br>**mothers** | **Mild-moderately**<br>**symptomatic mothers** | **Severe-critically**<br>**symptomatic mothers** |
| --- | --- | --- | --- |
|  | **59** | **78** | **8** |
| **Time between maternal infection and**<br>**delivery** |  |  |  |
| <60 days, n | 50 | 46 | 3 |
| 60-180 days, n | 6 | 26 | 5 |
| >180 days, n | 3 | 6 | 0 |
| **Trimester at the time of maternal**<br>**infection** |  |  |  |
| First Trimester, n | 3 | 8 | 0 |
| Second Trimester, n | 7 | 12 | 0 |
| Third Trimester, n | 49 | 58 | 8 |
| **Trimester at the time of delivery** |  |  |  |
| First Trimester, n | 0 | 0 | 0 |
| Second Trimester, n | 0 | 0 | 0 |
| Third Trimester, n | 59 | 78 | 8 |

**Supplemental Table 4:**
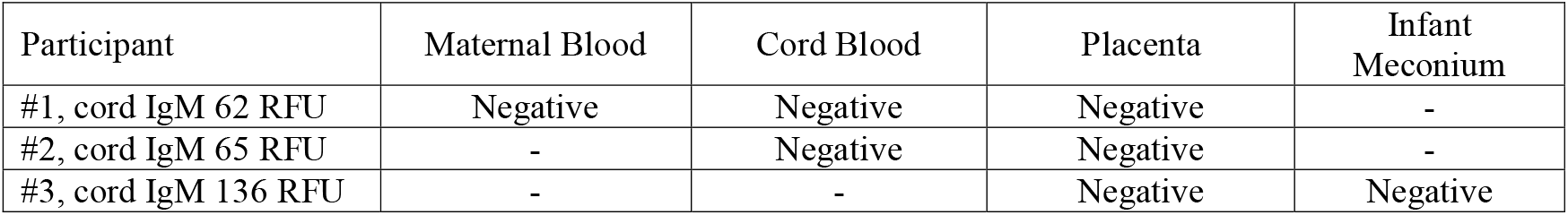
Delivery specimen PCR results.

| **Participant** | **Maternal Blood** | **Cord Blood** | **Placenta** | **Infant**<br>**Meconium** |
| --- | --- | --- | --- | --- |
| #1, cord IgM 62 RFU | Negative | Negative | Negative | - |
| #2, cord IgM 65 RFU | - | Negative | Negative | - |
| #3, cord IgM 136 RFU | - | - | Negative | Negative |

